# Long-term left atrial function after transcatheter device closure of patent foramen ovale in patients with cryptogenic stroke

**DOI:** 10.1101/2024.11.04.24316729

**Authors:** Teresa Gil-Jiménez, Rocía García-Orta, Inmaculada Villegas-Rodriguez, Juan Caballero-Borrego, Eduardo Moreno-Escobar

## Abstract

**Aims:** The implantation of percutaneous atrial septal occluder (ASO) devices for patent foramen ovale (PFO) may affect left atrial (LA) function. Some studies have reported short- to mid-term impairment of this function, though it remains unclear whether this is a transient or permanent negative effect, nor if all patients are equally susceptible to these changes.

**Methods and results:** Observational, prospective study of consecutive patients with cryptogenic stroke (CS) undergoing percutaneous closure of PFO. Echocardiographic evaluation of evolving structural and functional parameters of the LA was carried out before procedure, inmediately after (within 24 hours), 6 and 18 months after device insertion.

From March 2019 to October 2023, 170 patients with CS were enrolled, 82 eligible for PFO closure and 65 undergoing percutaneous closure. Baseline structural and functional parameters were within normal limits. At 6 months follow-up, there is a significant increase in LA volumes and a deterioration of reservoir and conduit functions measured by strain. There is a subsequent trend toward normalization, but baseline parameters are not reached by 18 months.

**Conclusions:** PFO device closure negatively impacts on LA function, resulting in increased atrial volumes, worsening of left ventricular diastolic function, decreased reservoir and conduit function, and a compensatory increase in pump function. These changes are significant at 6 months, with partial improvement but without full normalization of parameters at 18 months follow-up.

## INTRODUCTION

Patent foramen ovale (PFO) is a common finding in general population (20-25%). However, this feature is more frequently observed in young patients with a cryptogenic stroke (CS)^1,2^. Currently, scientific evidence^3^ suggests that percutaneous closure of the PFO in selected patients is more effective than antiplatelet therapy alone in preventing stroke recurrence. However, the incidence of atrial fibrillation (AF) after atrial septal occluder (ASO) implantation is the main adverse event^4^, resulting in a similar net clinical benefit between the two strategies. Even in selected patients, the superiority of device closure remains unclear^5,6^.

The percutaneous closure of atrial septal defect (ASD) or PFO is a very widespread interventional procedure, not only due to its efficacy and safety but also because it covers several pathologies and can be performed across a wide age range.

Several studies have evaluated the impact of these device implants on LA function, obtaining disparate results:

- On one hand, some studies show improvement in LA function parameters after the procedure, with normalization of pressures and reduction of pre-existing atrial dilation; this leads to a decreased incidence of supraventricular arrhythmias^7,8^. These patients, who experience improvement in atrial parameters, usually present some degree of prior “atrial myopathy”, secondary to their underlying condition (ASD, PFO with ASA, patients with respiratory pathology, or long-standing hypertension, etc.).
- On the other hand, most studies carried out in patients without atrial myopathy, such as those currently indicated for PFO closure after CS, demostrate that device implantation results in deterioration of atrial function parameters, though there are no definitive results regarding whether this effect is transient or permanent, or if all patients have the same vulnerability (studies with few patients and short- to medium-term follow-up)^9-14^.

Echocardiographic evaluation of LA function through volumetric measurements and speckle tracking techniques has proven to have both diagnostic and prognostic significance for predicting cardiovascular events across various clinical contexts^15,16^. Typically, strain during reservoir phase (peak atrial longitudinal strain at end systole) is the most significant parameter, and values below certain thresholds have been established in some conditions as predictive of the risk of AF onset or recurrence during follow-up^17-19^.

Our study evaluates LA function in patients with CS associated with PFO undergoing percutaneous closure. The evolution of reservoir, conduit, and pump functions is analyzed through phase-specific volumetric measurements and LA strain, with long-term follow-up.

## METHODS

### Study design

This observational, prospective study included all patients who experienced an ischemic stroke between March 2019 and October 2023, which, after a comprehensive diagnostic process, was classified as cryptogenic and associated with PFO. In acordance with the recommendations from the European Association of Percutaneous Cardiovascular Interventions (EAPCI)^20^, a multidisciplinary Heart-Stroke Team (HST) was established, consisting of neurologists, imaging cardiologists, and interventional cardiologists. A rigorous selection process was conducted to identify patients who met the criteria for PFO closure.

Clinical characteristics were collected for all patients, including sex, age, weight, height, the presence of classic cardiovascular risk factors, history of migraine, peripheral venous vascular disease, and thrombophilias), and the RoPE (Risk of Paradoxical Embolism) score was calculated. In all cases, a transthoracic echocardiogram was performed (measuring phase-specific LA volumes and LA strain parameters), along with agitated saline contrast and transesophageal echocardiography (assessing PFO characteristics), and the PASCAL score (PFO-Associated Stroke Causal Likelihood) was also calculated.

In patients undergoing percutaneous PFO closure, procedural characteristics such as the size of the device (Figulla® Flex II PFO Occluder) and residual shunt were taken into account. A control echocardiogram was performed after procedure at 24 hours, as well as at 6 and 18 months during follow-up, utilizing both transthoracic and agitated saline contrast. All patients underwent clinical follow-up and a 24-hour Holter monitoring at 6 and 18 months.

The study was conducted in accordance with the guidelines of the Declaration of Helsinki and approved by the Ethics Commitee of Granada’s Hospital Universitario Clínico San Cecilio (No. 0372-N-19), receiving approval on 5 th March 2019. Informed consent was obtained from all subjects involved in the study.

### Echocardiographic asessment of LA function^21-23^

Transthoracic and transesophageal echocardiographic images were performed using Vivid E95; GE Medical Systems; equiped with 4Vc probe (2-4MHz) transducers. These images were subsequently digitized and post-processes offline by the same experienced operator using the EchoPAC workstation.

LA function was evaluated using volumetric measuring and two-dimensional speckle tracking echocardiography (2D STE). LA volume was assessed using the biplane method of disks, tracing the LA inner border, and excluding the area under the mitral valve annulus, the LA appendage and the inlet of the pulmonary veins. The maximal, minimal and pre-A LA volumes were mesured just before the opening and at the closure of the mitral valve, and at the onset of P-wave on the ECG, respectively. All volumes were indexed to the body surface area and expressed in millilitres/metres squared. LA function indexes were calculated using the measured values. The formulas used are as follows:

- LA reservoir function was characterized by using: LA total emptying volume = (Vmax – Vmin) and LA total emptying fraction = [ (Vmax − Vmin)/Vmax]x100, LA expansion index = [ (Vmax − Vmin)/Vmin]x100.
- LA conduict function using LA passive emptying volume = (Vmax − Vpre-A), LA passive emptying fraction = [(Vmax − Vpre-A)/Vmax]x100.
- LA contraction function using LA active emptying volume = (Vpre-A – Vmin), LA active emptying fraction = [ (Vpre-A − Vmin)/Vpre-A]x100.

LA myocardial strain was evaluated using the same software (EchoPAC; GE Medical Systems). We analyzed the lateral and septal wall from the apical 4-chamber view, and the anterior and inferior wall from the apical 2-chamber view and calculated the average of the strain curves. Zero strain was set at the QRS onset. In the strain curve we evaluated *(Figure 1):*

**Figure 1:**
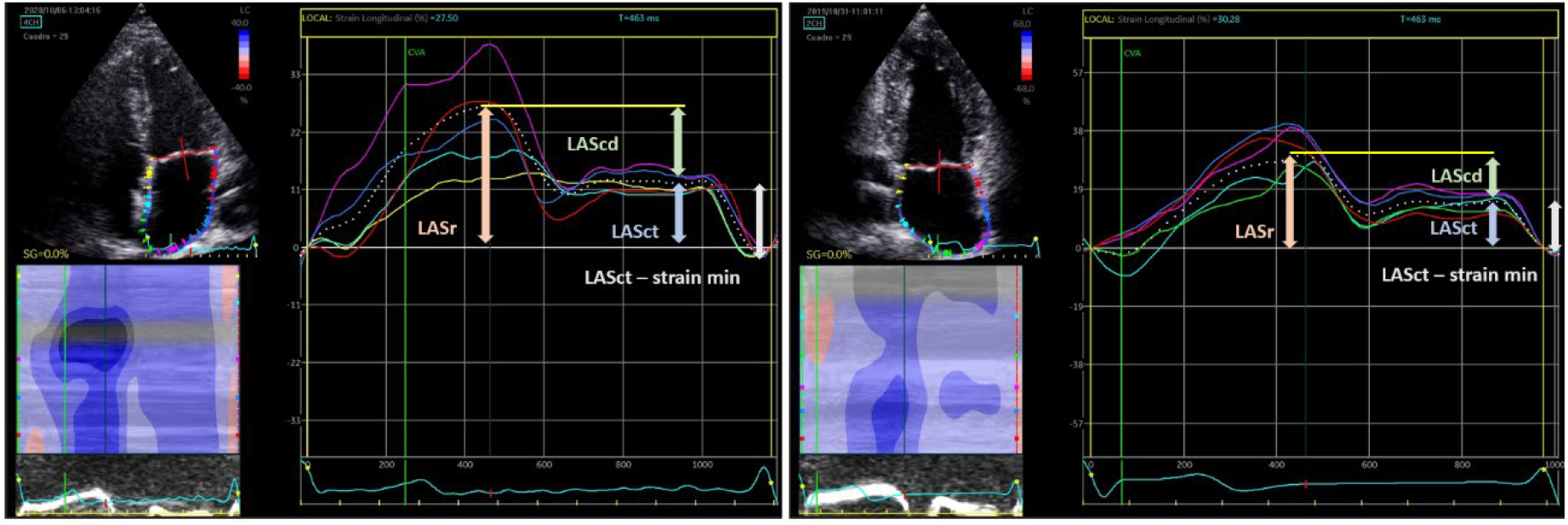
LA speckle tracking analysis using EchoPAC; GE Medical Systems.

- LA reservoir function (LASr): Peak systolic strain at the end of ventricular systole.
- LA contraction function: Peak in early diastolic strain before atrial contraction (LASct) and LA passive emptying strain (LASct-minimal strain).
- LA conduict function (LAScd): passive emptying strain (LASr-LASct).

### Statistics

A specific data collection form was designed, and the statistical analyses were performed using IBM SPSS version 25. First, data coding and exploration were conducted. Second, a descriptive analysis of the set of variables was performed, using frequencies and percentages for categorical variables and mean and standard deviation for quantitative variables. Chi-square tests were used to study the relationship between variables, and Student’s t-test for independent samples was applied to determine if there were differences between groups. Additionally, a repeated measures ANOVA was conducted to assess whether changes occurred over time. In all cases, a 95% confidence level was used.

## RESULTS

Of the 170 patients with CS analyzed, 82 patients were selected as candidates for percutaneous PFO closure based on current recommendations (59.8% male, mean age 47.12 years, mean RoPE score 6.66, and 39% classified as “probable” on the PASCAL scale). Of these 82 patients with an indication for PFO closure, the procedure was performed in 65 of them *(Figure 2)*.

**Figure 2:**
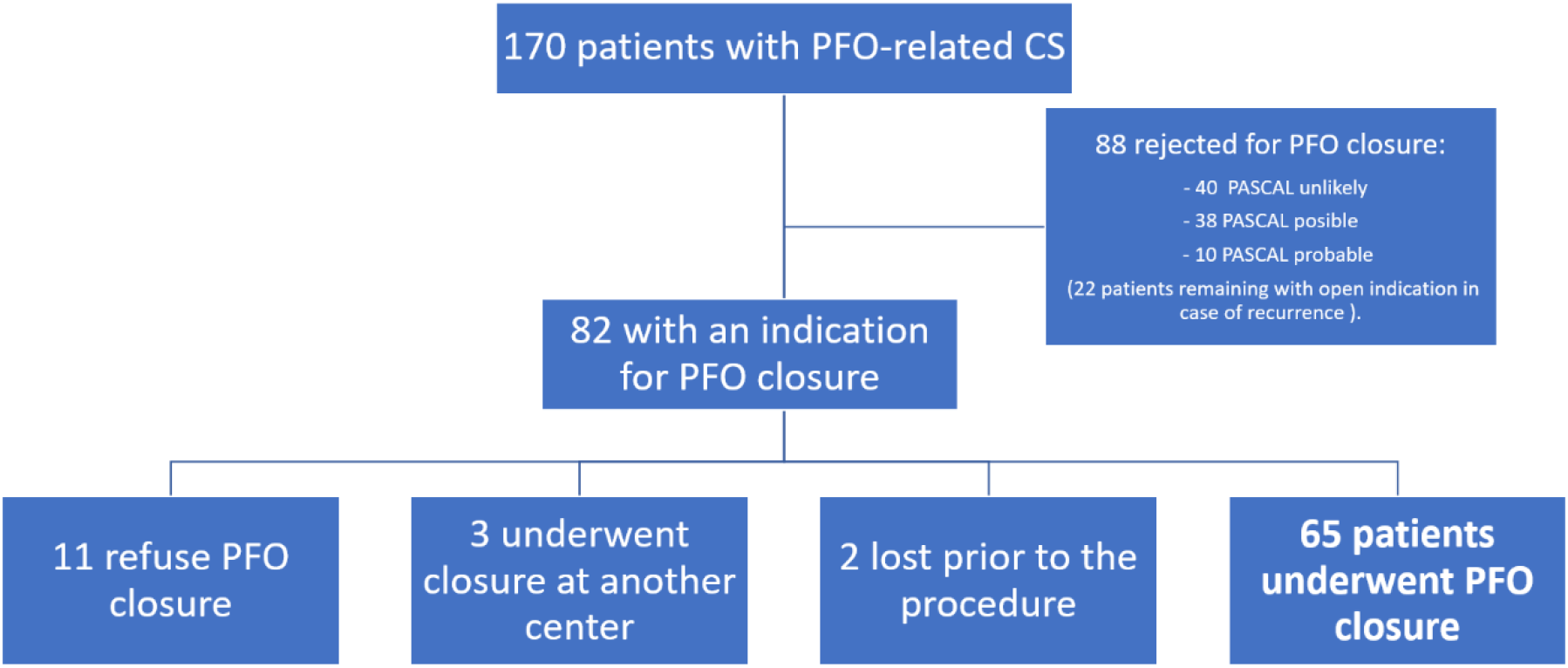
Flowchart illustrating enrolment process of study participants.

**Figure 3:**
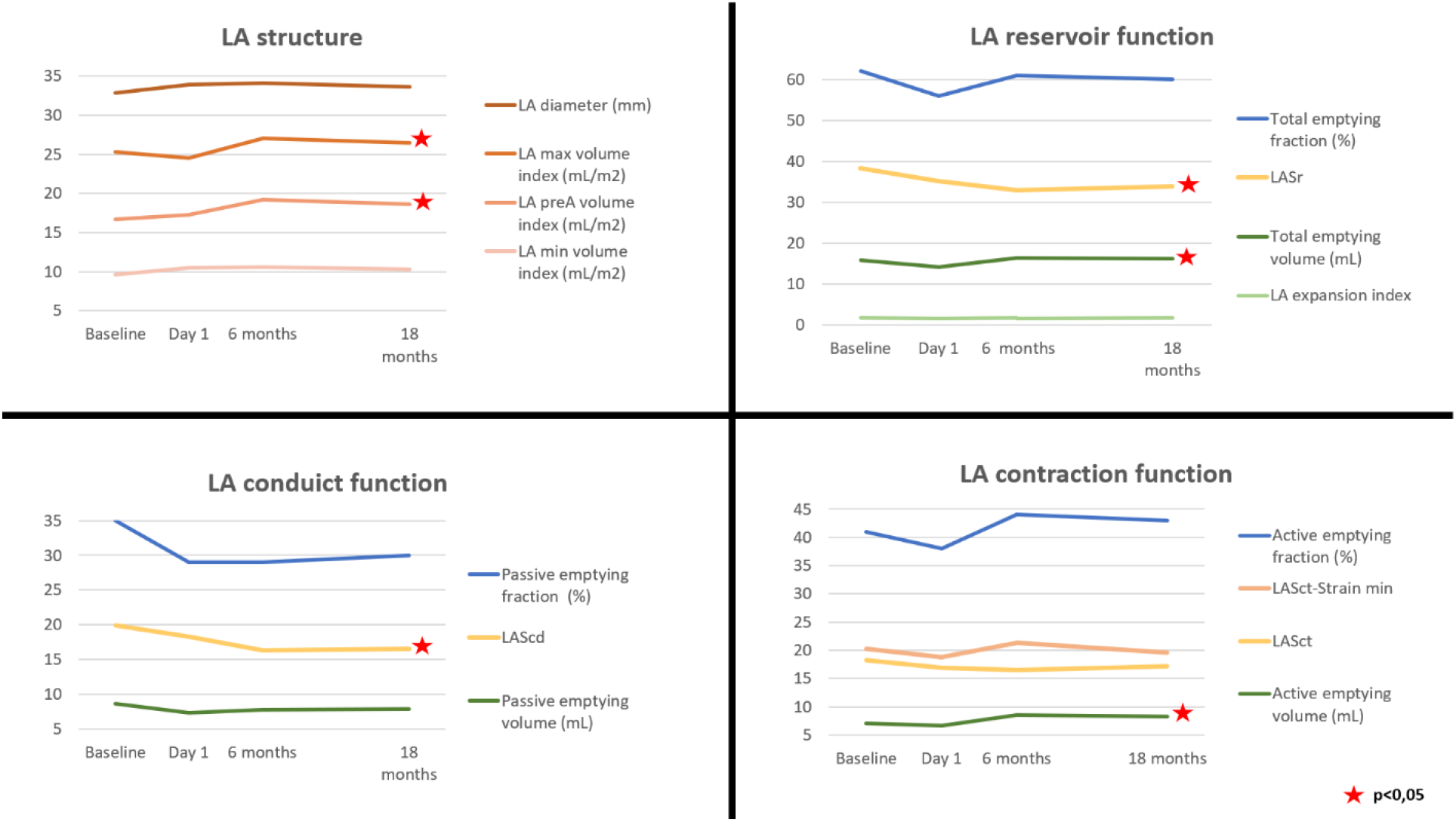
Evolution in monitoring structural and functional and echocardiographic parameters. After PFO closure, the immediate assessment at 24 hours post-procedure revealed a non-significant reduction in the maximum LA volume. However, at 6 months, there was a significant dilation of the phase-specific volumes (maximum and pre-A), which subsequently showed a tendency to normalize, though baseline volumes were not fully restored at 18 months. This dilation, more pronounced at 6 months, was associated with a significant increase in total and active emptying volumes, alongside a non-significant increase in passive LA emptying volume.

*Table 1* show the baseline characteristics of patients who underwent PFO closure. There was a slight predominance of males, with an average age around 47 years, a low prevalence of classic cardiovascular risk factors, but 43.1% are current or former smokers, and the overall body mass index indicates overweight. The RoPE score was over 6, and the PASCAL score was classified as “possible” or “probable” in most cases.

**Table 1:** Baseline clinical and conventional echocardiographic characteristics.

| Table 1: Baseline clinical and conventional echocardiographic characteristics | 65 patients |
| --- | --- |
| Age (years) | 46,82 |
| Gender (male) | 56,9% |
| Hypertension | 24,6% |
| Diabetes Mellitus | 6,2% |
| Hypercholesterolemia | 24,6% |
| Former + current smoker | 43,1% |
| Body mass index (kg/m2) | 26,05 |
| No trombophilias | 73,8% |
| History of stroke/TIA | 6,2% |
| Cortical infarct on imaging | 81,5% |
| Peripheral venous vascular disease | 3,1% |
| History of migraine | 20% |
| RoPE score | 6,72 (3-10) |
| PASCAL score | Probable 41,5%<br>Possible 52,3% |
| ASA | 27,7% |
| Prominent Eustachian valve or Chiari's network | 18,5% |
| Shunt at rest/Valsalva maneuver | Large 24,6%→63,1% |
| E/A ratio | 1,12 |
| LVEF (%) | 59,81 |

**Table 2:** Baseline structural and functional echocardiographic characteristics. Regarding baseline echocardiographic characteristics, all parameters were within normal limits according to the standards set by the American Society of Echocardiography and the European Association of Cardiovascular Imaging^21-23^.

| Table 2: Baseline structural and functional echocardiographic characteristics | 65 patients |
| --- | --- |
| **LA structure** |  |
| LA diameter (mm) | 33,85 |
| LA maximum volume index mL/m2 | 26,37 |
| LA pre-A volume index mL/m2 | 17,12 |
| LA minimum volume index mL/m2 | 9,58 |
| **LA reservoir function** |  |
| Total emptying volume (mL) | 16,79 |
| Total emptying fraction (%) | 63,5 |
| LA expansion index | 1,92 |
| LASr | 38,08 |
| **LA conduit function** |  |
| Passive emptying volume (mL) | 9,25 |
| Passive emptying fraction (%) | 35,5 |
| LAScd (LASr-LASct) | 19,01 |
| **LA contraction function** |  |
| Active emptying volume (mL) | 7,54 |
| Active emptying fraction (%) | 43,05 |
| LASct | 19,07 |
| LASct-minimal strain | 21,10 |

As reported in other studies, this atrial dilation was linked with a significant deterioration in longitudinal strain parameters, particularly affecting reservoir (LASr) and conduit function (LAScd). Similar to the volumen changes, this decline in parameters was significant at 6 months, with a trend toward normalization by 18 months.

No differences in atrial function performance were observed based on ASO size or residual shunt persistence. However, a different response in atrial function was observed in subjects with atrial septal aneurysm (ASA); these patients (n=18, 27.7%) initially presented lower atrial strain values and experienced less deterioration in these parameters after device implantation *(Figure 4)*.

**Figure 4:**
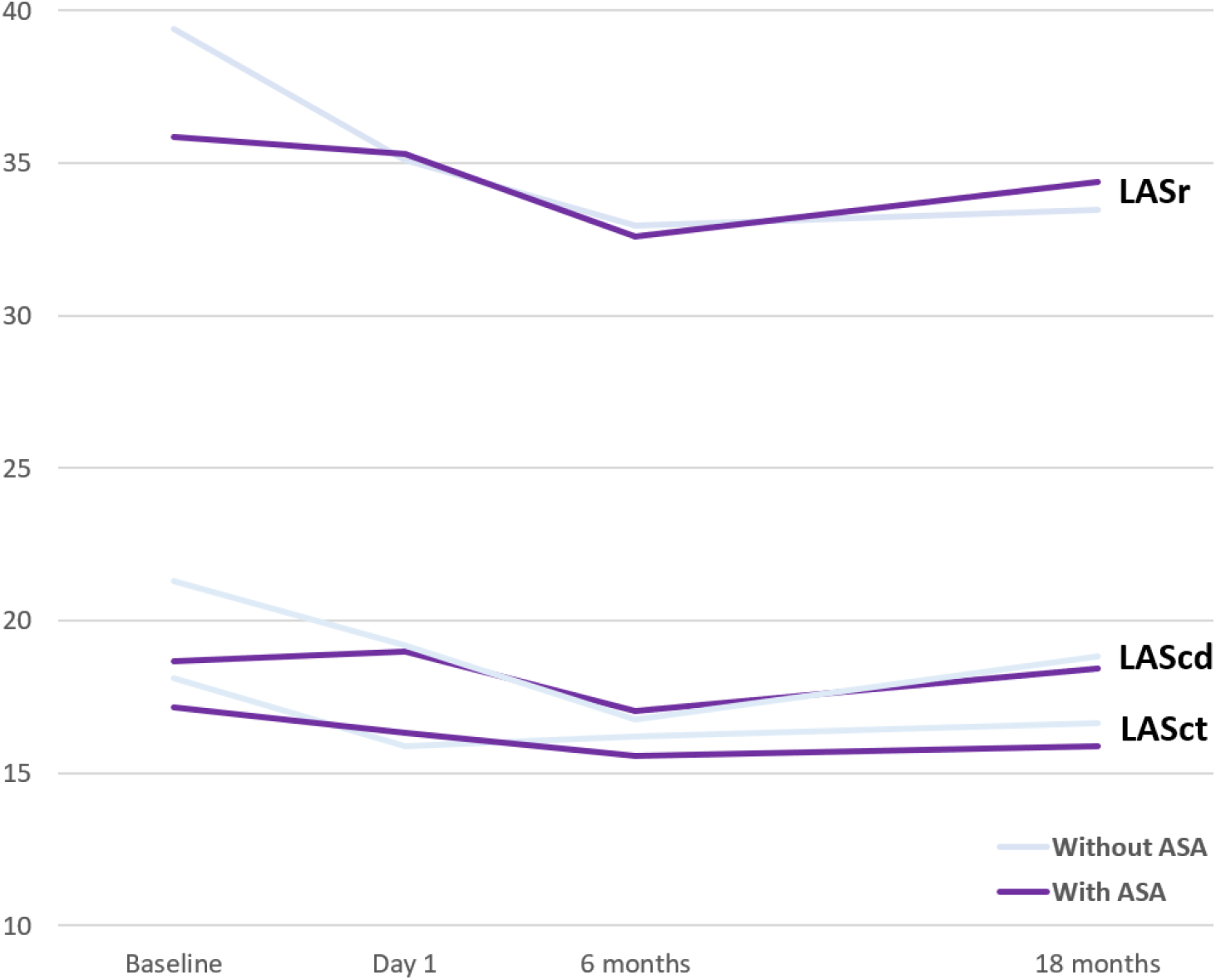
Comparison of strain evolution in patients With and Without ASA

Likewise, an analisis of the subgroup consisting of 28 patients who initially exhibited lower longitudinal strain parameters (specifically, LASr below the average) revealed that these patients experienced a non-significant improvement in reservoir and conduit strain parameters (LASr and LAScd) within 24 hour following immediate closure. The decline at 6 months was almost imperceptible, and there was a complete recovery by 18 months *(Figure 5)*.

**Figure 5:**
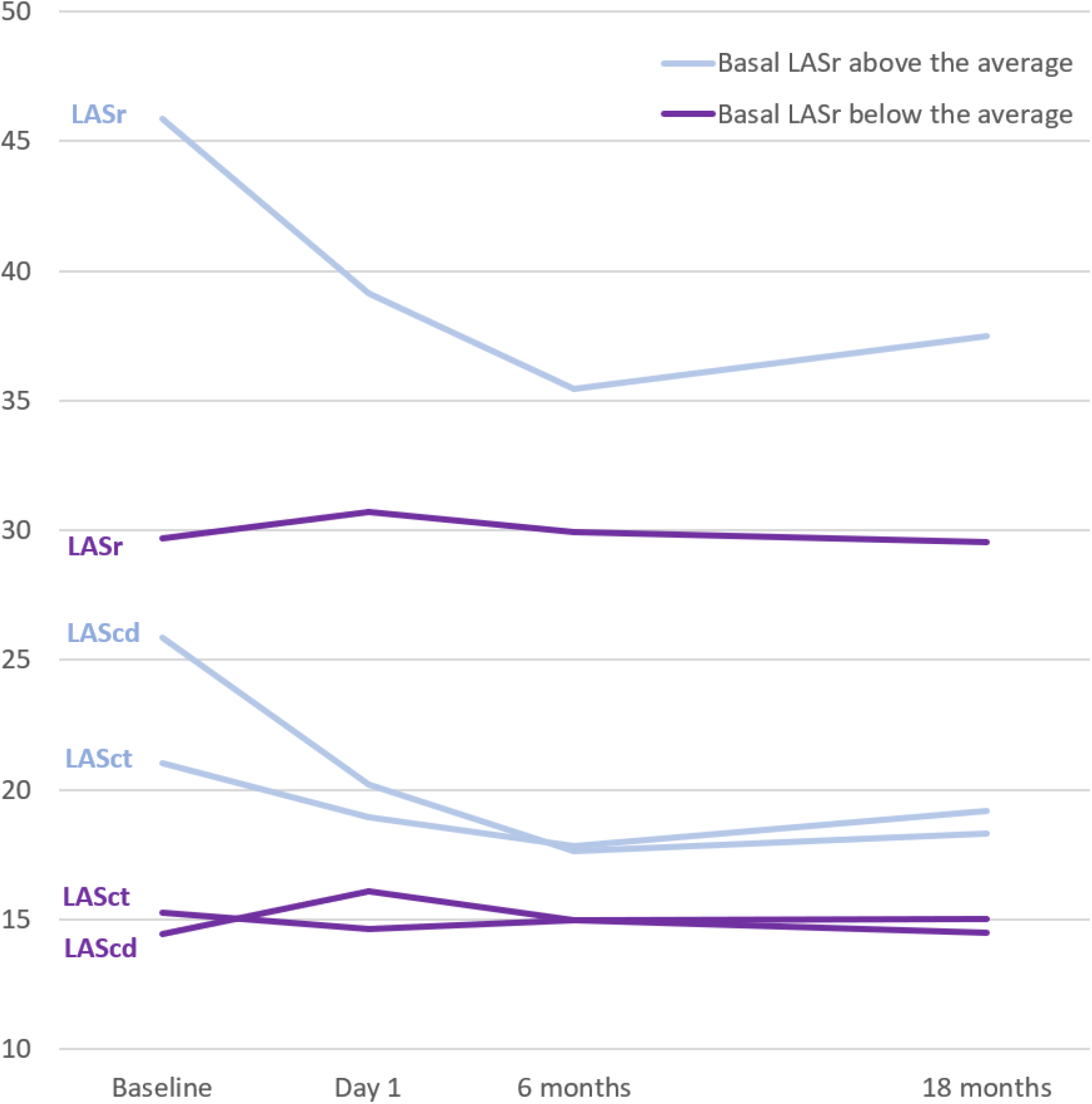
Comparison of strain evolution in patients with basal LASr above and below the average.

During the clinical follow-up of patients undergoing PFO closure (which, in the earliest cases, has extended beyond 5 years), 84.6% have remained event-free. AF/flutter occurred in 3 patients (4.6%) within the first month following the closure; 2 patients (3.1%) experienced recurrent ischemic stroke, for which empirical anticoagulation was initiated; and 5 patients (7.7%) had transient ischemic attacks (TIA) without new lesions on brain imaging. An analysis of atrial function in these patients (especially in those who developed AF) did not reveal any statistically significant differences.

## DISCUSSION

In our study, the implantation of a ASO in patients with PFO related CS and normal baseline LA function resulted in a significant deterioration of atrial functional parameters and strain at 6 months. This was followed by improvement, although full normalization was not achieved at 18 months. In contrats, patients with reduced baseline atrial function experienced either a less pronounced impairment or an imperceptible change.

Percutaneous PFO closure is a widely performed procedure. Over the years, the evidence supporting its use for prevention of stroke recurrence has evolved, shaped by various clinical trials. Currently, candidates who benefit from ASO implantation are strictly selected: they are tipically under 60 years of age, exhibit a low burden of cardiovascular risk factors (RoPE score greater than 6), and present a PFO with high-risk features, making the PASCAL score “probable” or “highly probable” ^24^. These patients generally do not have structural or functional heart disease (except for the presence of PFO).

Device implantation for ASD closure has become the standard therapeutic strategy over surgical options, but the negative impact of this prosthetic material on “healthy” hearts remains a subject of ongoing study and reevaluation^25^, even leading to the withdrawal of models associated with a higher incidence of AF after the procedure.

In addition to classic studies on residual shunt rates and supraventricular arrhythmias following ASO implantation^26^, there are more advanced studies evaluating LA function after the procedure:

- In 13 patients, percutaneous ASD closure resulted in significant global and segmental atrial dysfunction, measured by deformation parameters, which persisted at 6 months post-implantation^9^.
- In 25 patients after PFO closure, segmental strain of the LA anterior wall decreased at 3 months but was compensated by an increase in lateral wall strain, resolving at 6 months. Non-significant changes in atrial volumes (increased maximun volumen) maintained at 6 months^10^.
- In a study comparing healthy patients to those undergoing either percutaneous or surgical ASD closure, at 1 year post-intervention, the group that received percutaneous device closure (n=20) showed reduced LA reservoir strain, conduit function, and contraction function compared to healthy controls (n=20), while there were no differences with the surgical group (n=42)^12^.
- A retrospective study including 39 patients who underwent percutaneous PFO/ASD closure found that reservoir parameters remained significantly decreased at 1 year post-implant (LASr 32.8 ± 13.9% vs. 26.7 ± 10.7%, p = 0.01)^13^.
- A comparative study of PFO closure with device implantation (n=20) vs. suture (n=20) showed that the novel suture technique, which leaves no metallic material in the septum, did not affect LA function, unlike the device, which impaired it, potentially leading to arrhythmias^14^.

Our study prospectively evaluated 65 carefully selected patients with CS and an indication for PFO closure. We followed these patients for 18 months after the procedure. Our findings corroborate previous research, revealing that implantation significantly increased atrial volumes and impaired atrial function, as measured by 2DSTE techniques. The most pronounced deterioration occurred at 6 months, followed by gradual improvement; however baseline values were not reached at 18 months.

Nearly all patients had baseline echocardiographic parameters within normal ranges. However, those with initially lower LASr values showed a different response. At 24 hours post-procedure, they experienced a slight improvement in reservoir and conduit function, with an almost minimal decline at 6 months and complete recovery at 18 months. These patients likely presented with an initial form of “atriopathy,” suggesting that the impact of the device on atrial function was less significant, though this cannot be confirmed due to the small patient sample.

Patients with ASA fall within this group of individuals exhibiting baseline LASr values below the average (at the lower limit of normal), which potentially indicative of early signs of atriopathy. After device implantation, they experienced less deterioration in LA function. This reduced impairment, combined with the fact that ASA is associated with a higher embolic risk, makes this population particularly favorable for this therapy.

Given that the benefits derived from the implantation of PFO closure device in these selected patients still come with complications, particularly post-implant supraventricular arrhythmias, the indications for the procedure and the ASOs used must continue to evolve to minimize them. Medium- and long-term studies assesing atrial function in these patients could provide valuable insignts into to the impact of closures. Smaller, more flexible, or those resembling surgical suture techniques^14^ may enhance atrial functional parameters and help us move closer to the goal of zero complications.

Long-term studies involving a larger cohort of patients are needed to determine whether the effect of the device diminish or disappear over time, or, conversely, if they persist.

The limitations of our study arise mainly from the small patients sample, which may lead to insufficient power to evaluate outcomes at 18 months. This also precludes precise subgroup analysis, for example, of patients with or without atriopathy, or ASA, etc. Additionally, this study focuses on patients at high risk of CS secondary to PFO, according to risk scales and a strict consensus between the cardiology-neurology team; therefore does not apply to patients with higher cardiovascular comorbidity who undergo this procedure.

## Data Availability

All data included in the study are available in a database specifically created for this research.

## Acknowledgments

None.

